# COVID-19 mortality prediction model, 3C-M, built for use in resource limited settings - understanding the relevance of neutrophilic leukocytosis in predicting disease severity and mortality

**DOI:** 10.1101/2021.08.05.21261565

**Authors:** Niharika Agarwal, Devika Dua, Ritika Sud, Madhur Yadav, Aparna Agarwal, Vijesh Vijayan

## Abstract

In this study, a combination of clinical and hematological information, collected on day of presentation to the hospital with pneumonia, was evaluated for its ability to predict severity and mortality outcomes in COVID-19. Ours is a retrospective, observational study of 203 hospitalized COVID-19 patients. All of them were confirmed RT-PCR positive cases. We used simple hematological parameters (total leukocyte count, absolute neutrophil count, absolute lymphocyte count, neutrophil to lymphocyte ration and platelet to lymphocyte ratio); and a severity classification of pneumonia (mild, moderate and severe) based on a single clinical parameter, the percentage saturation of oxygen at room air, to predict the outcome in these cases. The results show that a high absolute neutrophil count on day of onset of pneumonia symptoms correlated strongly with both severity and survival in COVID-19. In addition, it was the primary driver of an initial high neutrophil-to-lymphocyte ratio (NLR) observed in patients with severe disease. The effect of low lymphocyte count was not found to be very significant in our cohort. Multivariate logistic regression was done using Python 3.7 to assess whether these parameters can adequately predict survival. We found that clinical severity and a high neutrophil count on day of presentation of pneumonia symptoms could predict the outcome with 86% precision. This model is undergoing further evaluation at our centre for validation using data collected during the second wave of COVID-19. We present the relevance of an elevated neutrophil count in COVID-19 pneumonia and review the advances in research which focus on neutrophils as an important effector cell of COVID-19 inflammation.

## Introduction

The COVID-19 pandemic is caused by the severe acute respiratory syndrome coronavirus 2 (SARS CoV-2) infection(1), (2). The illness has a wide range of presentations ranging from a completely asymptomatic presentation to life threatening lung involvement(3), (4), (5), (6). The COVID-19 pneumonia is a lung inflammation syndrome which is triggered by a cytokine storm(7), (8), (9), (10). It is important to note that ‘Cytokine Storm Syndrome’ is not a precisely defined entity and there is no diagnostic test/ criteria validated for accurately diagnosing it in the context of COVID. However, several inflammatory markers have been identified as markers of severity for this illness with a special emphasis on IL-6(11), (12). Our understanding of this disease is evolving in a dynamic fashion as more data becomes available for study and every fresh piece of information points towards newer directions.

Since the beginning efforts have been made to identify parameters which could be indicative of severity and mortality. One of the most persistent findings, replicated in many studies, has been the association of lymphopaenia, as well as an elevated NLR, with severity and mortality in COVID(13), (14), (15), (16)(17). Other commonly used markers to predict adverse outcomes include presence of elevated D-Dimer levels indicative of a hyper-coaguable state, elevated Ferritin, Procalcitonin and IL-6 levels; and CT severity(17), (18), (19), (20), (21), (22).

A great drawback is that this information is not routinely available especially in a primary health care setting which is frequently the first point of contact for patients. Some of the assays do not, as yet, have an international standard for reporting making it difficult to interpret the results with accuracy. Our aim was to evaluate easily available information at presentation in order to discern early markers of adverse outcomes. The markers which potentially have a wide applicability even in resource limited settings.

## Materials and Methods

This study is a retrospective analysis of 203 COVID-19 patients diagnosed at Lady Hardinge Medical College, Delhi. They were diagnosed based on the reverse transcription quantitative polymerase chain reaction (RT-qPCR) tests done on respiratory secretions obtained via nasal swabs.

Demographic, clinical and laboratory data was collected at the time of presentation to the hospitals with respiratory symptoms. For our study we considered a single, easily available clinical parameter, the percentage oxygen saturation (SpO2) at room air as measured by a pulse oximeter. The patients were a assigned a clinical severity class on the basis of this variable. Those with SpO2 >/= 95% were considered mild, those with SpO2 between 90-94% were considered moderate, and those with SpO2 < 90% were considered severe. A complete blood count done in accordance with laboratory standards, on the day of presentation to the hospital [total leukocyte count, absolute neutrophil count, absolute lymphocyte count, platelet count, hemoglobin, neutrophil to lymphocyte ratio and platelet to lymphocyte ratio], was considered.

This study was performed in line with the principles of the declaration of Helsinki after approval from the local ethics committee.

## Statistical Analyses

Multivariate analysis was conducted using Python 3.7. Tests used included the Pearson chi-square and ANOVA to compare the differences between two categorical variables. Pair wise Tukey test was used to calculate between class significance of a variable. Precision recall curve was used to generate an AUC for the prognostic variable under study. Multivariate logistic regression analysis in Python (ols model) was used for the computation of significance of multiple variables and their correlation with each other. A p-value below 0.05 was accepted as statistically significant. A predictive model was built using the log regression analysis in Python and is currently available for us on the internet.

## Results

Demographics - We retrospectively studied hematological investigation of 203 COVID 19 positive patients of which 58.62% (n=119/203) were males and 41.37% (n=84/203) were females. Mean age of study sample was 46 ± 16.37 years ranging from 19 years to 82 years. Out of 203 COVID 19 positive patient 152 (74.87%) survived and 51 (25.12%) succumbed. 90 patients had mild disease, 55 moderate and 58 patients had severe COVID19 disease.

All of the hematological data was analyzed to generate a correlation matrix (table 1) which would provide valuable information regarding the relationship between the various variables. It aided recognition of variables which are highly correlated (table 2) and helped reduce redundancy of data.

**Table 1&2:**
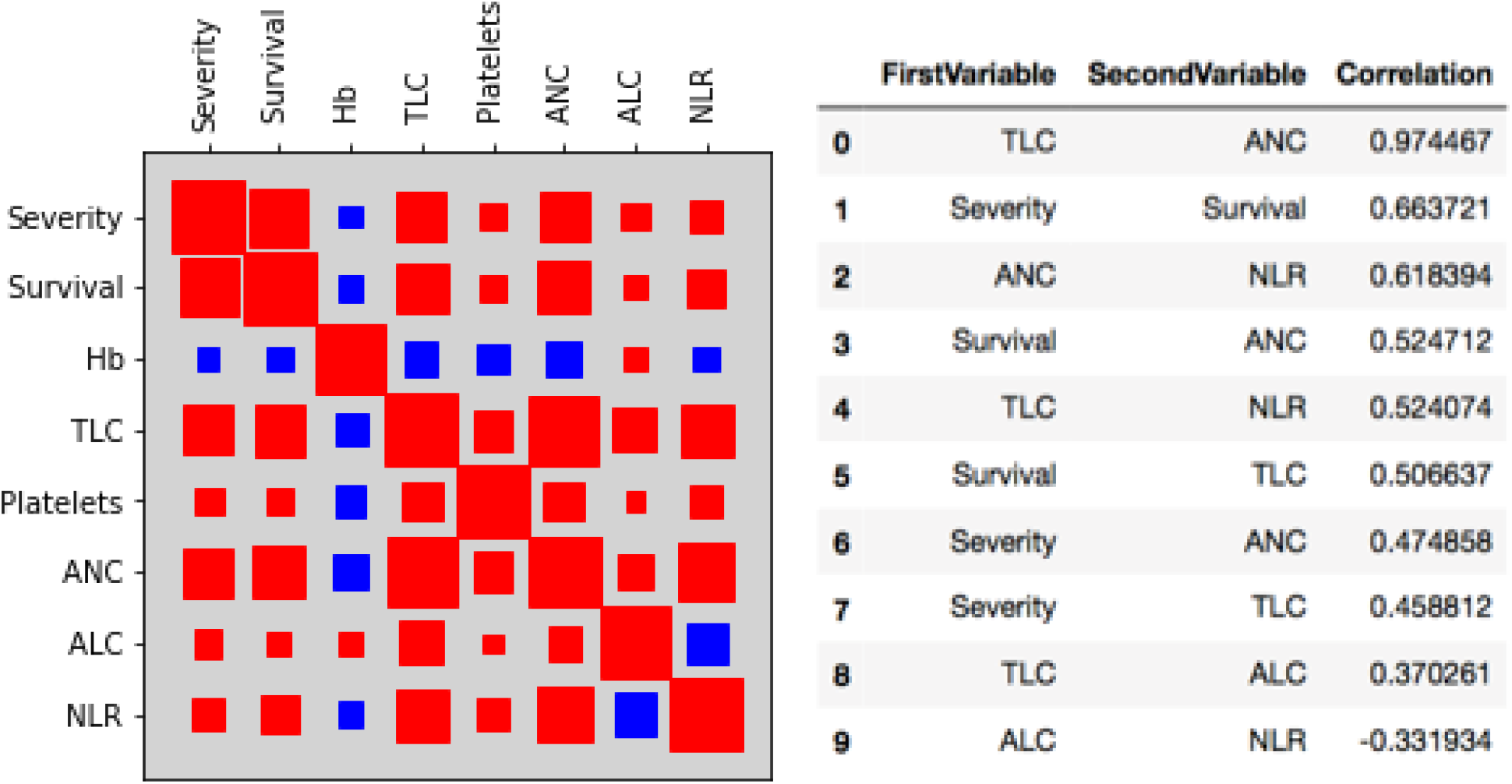
Correlation matrix showing the inter-relationship of variables under study

Of these variables, TLC-ANC, Survival-Severity, ANC-NLR, Survival-ANC, TLC-NLR and TLC-Survival were found to have significant positive correlation with one another. ALC was found to have a negative correlation with both NLR and survival but the effect size was not significant.

The difference between the mean of absolute neutrophil count among those who survived versus those who didn’t was compared using the pair wise Tukey test and was found to be highly significant with a p value of 0.001. ANC mean was also significantly different among those with mild, moderate and severe disease. The difference between mild-moderate and mild-severe categories was more significant with a p value of 0.001 whereas the difference between moderate-severe was slightly less with a p value of 0.005. Although the results were significantly different the generated ANC cut off had an AUC of 0.66 in a Precision-Recall curve (used for an imbalanced dataset) and therefore an absolute cut off value of ANC for predicting survival could not be generated with sufficient confidence.

**Table 3:**
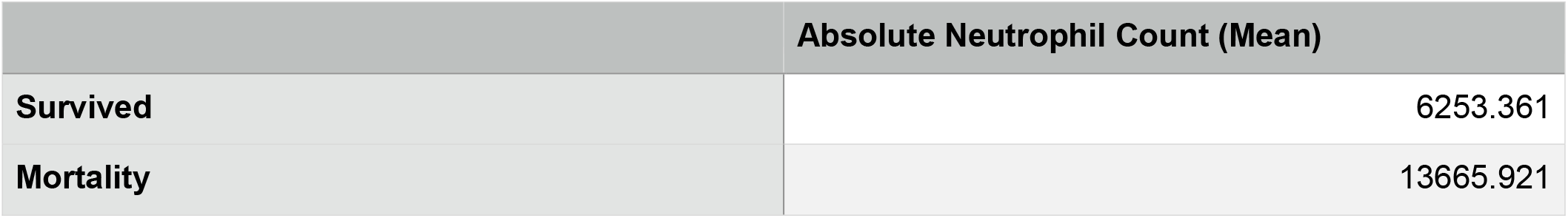
Mean ANC in those who survived versus those who did not

**Table 4:** Results comparing ANC in those who survived versus those who did not using p-tukey

| Category | diff | se | T | p-tukey | Hedges |
| --- | --- | --- | --- | --- | --- |
| Survival-Mortality | -7412.559 | 848.245 | -8.738 | 0.001 | -1.408 |

**Figure 1:**
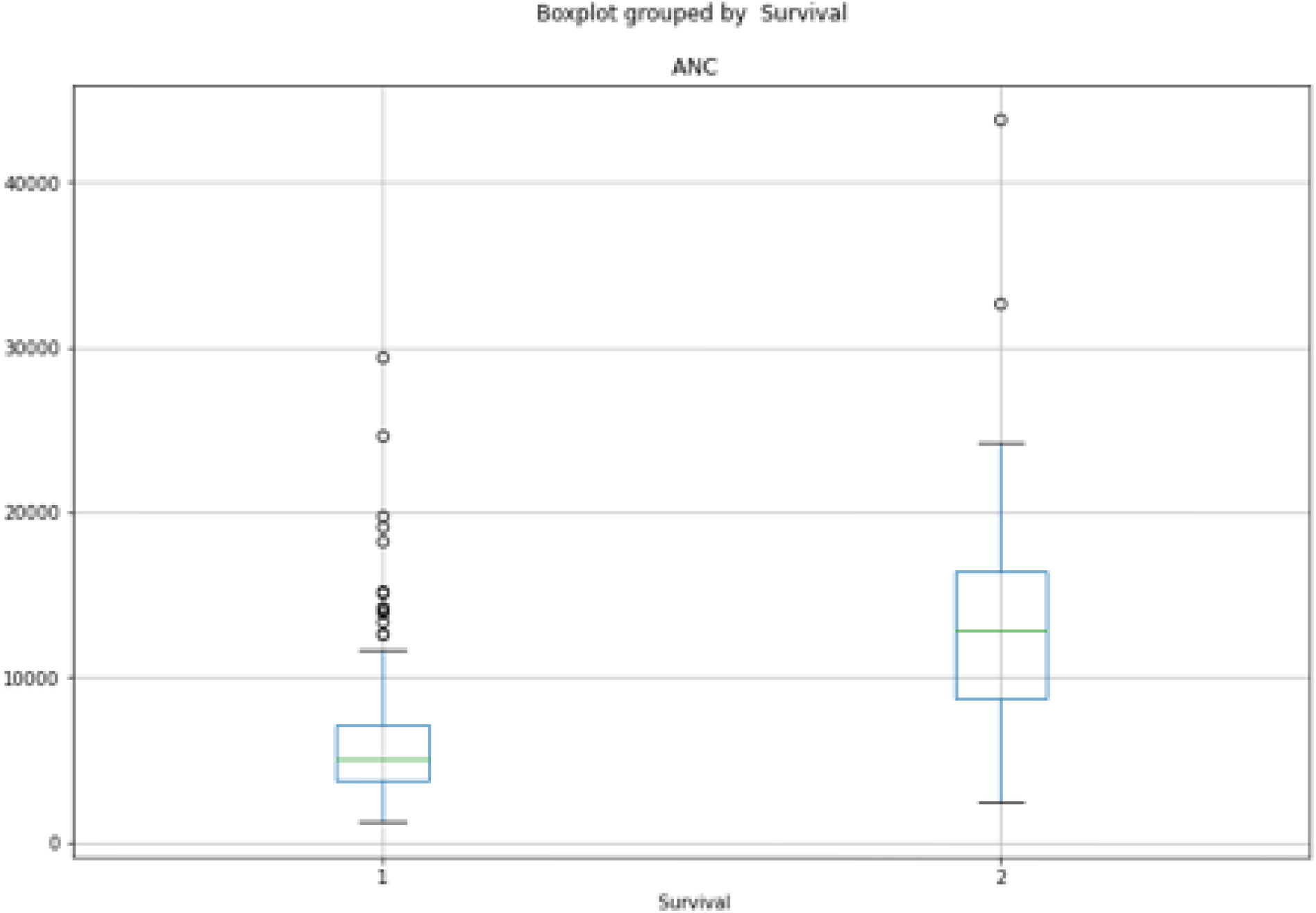
ANC boxplot grouped by survival

**Table 5:** Mean ANC in mild, moderate and severe categories of disease

| Severity | Absolute Neutrophil Count |
| --- | --- |
| Mild | 5146.911 |
| Moderate | 8819.709 |
| Severe | 12054.603 |

**Table 6:** Results comparing mean ANC with disease severity using p-tukey

| Category | diff | se | T | p-tukey | hedges |
| --- | --- | --- | --- | --- | --- |
| Mild-Moderate | -3672.797 | 929.642 | -3.950 | 0.001 | -0.672 |
| Mild-Severe | -6907.692 | 914.598 | -7.552 | 0.001 | -1.265 |
| Moderate-Severe | -3234.894 | 1022.300 | -3.164 | 0.005 | -0.591 |

**Figure 2:**
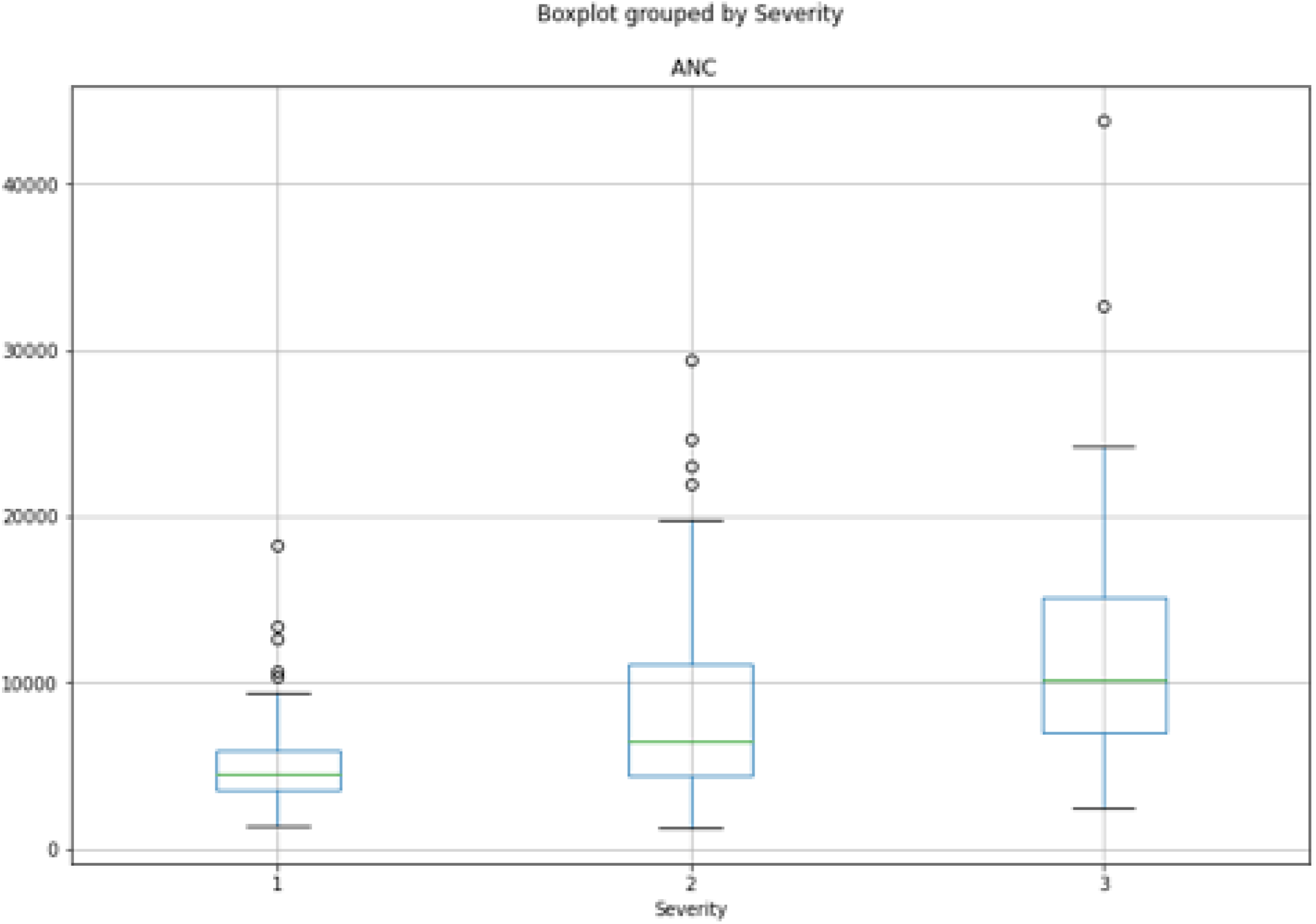
Boxplot of ANC grouped by severity

The absolute lymphocyte count was on the lower side in all severity classes but contrary to expectations the lowest value in our cohort was served in the mild category with progressive, minimal increase in the moderate and severe categories. The difference was not significantly associated with severity. Same held true for the comparison between those who survived with those who didn’t. The count was somewhat higher in the mortality group and there was no significant difference between the two. The overall incidence of lymphopaenia defined as absolute lymphocyte count < 1500/mL was 26.2% in our cohort which is similar to the findings in other studies but there was no correlation with severity or survival.

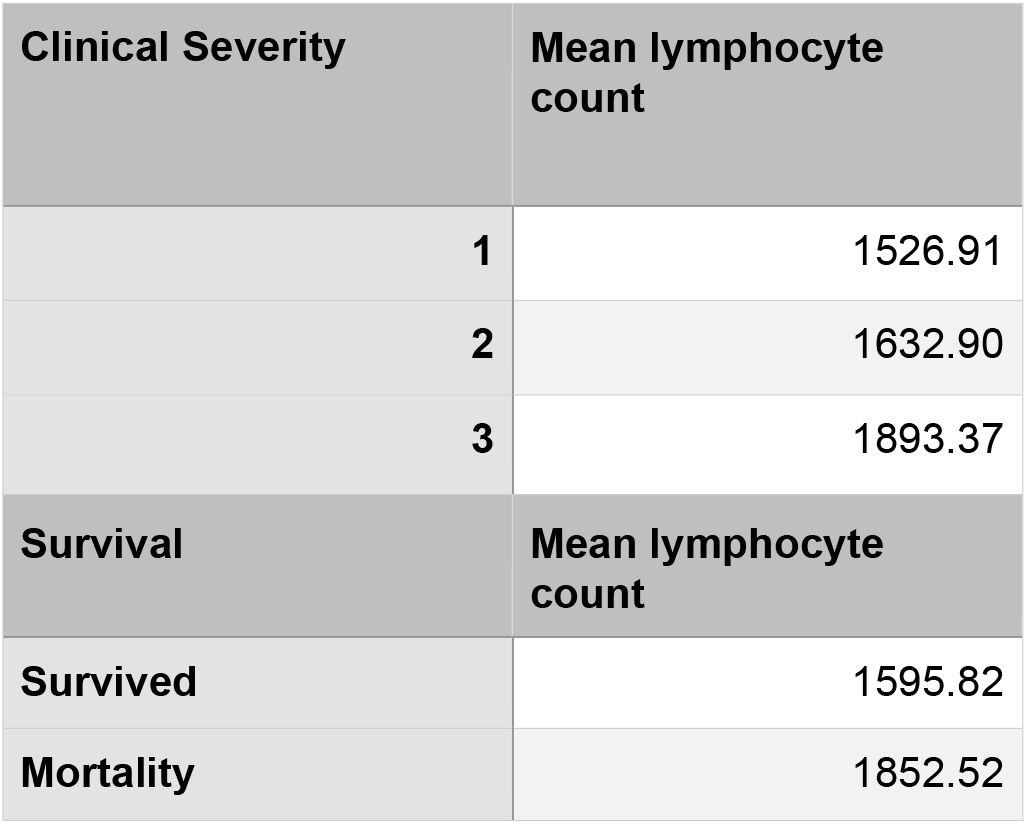
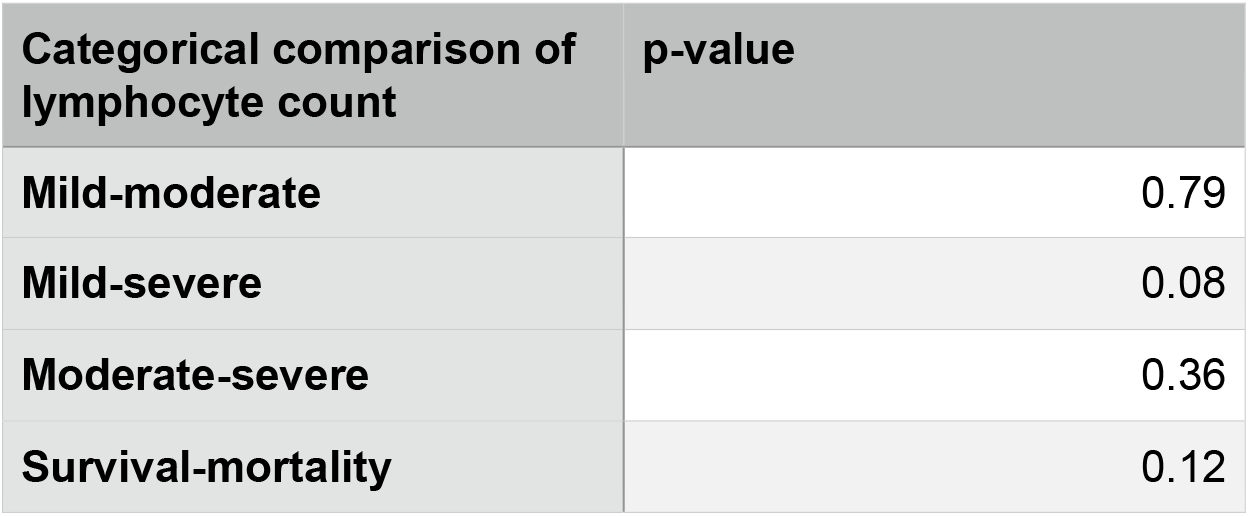

The NLR (neutrophil to lymphocyte ratio was also assessed independently for its association with severity and survival. The NLR was significantly different in the mild-severe categories and in the survival-mortality categories. But it’s important to note that NLR correlated greatly with absolute neutrophil count bringing an element of redundancy to its use as an independent predictor of survival and mortality in our cohort.

| Clinical severity scale | Mean NLR |
| --- | --- |
| Mild | 4.92 |
| Moderate | 7.76 |
| Severe | 10.00 |

| Survival | Mean NLR |
| --- | --- |
| Survived | 5.30 |
| Mortality | 12.56 |

| Categorical comparison of mean NLR | p-value |
| --- | --- |
| Mild-moderate | 0.27 |
| Mild-severe | 0.01 |
| Moderate-severe | 0.52 |
| Survival-mortality | 0.001 |

Multivariate logistic regression modeling was done in Python 3.7 using clinical severity, absolute neutrophil count, total leukocyte count, absolute lymphocyte count, NLR, PLR as assessed on day of presentation to hospital, to predict survival outcome of patient. The data was scaled prior to processing. The aim was to see if simple, point of care information obtained at the time of presentation could help guide initial triage and alert the physician on day 1 about those who would require more vigilant monitoring and care. The model had an overall accuracy of 87%. The parameters ANC, TLC and severity had significant association with mortality with p values approaching zero. Of these, ANC and TLC correlate very highly with one another as previously seen. ALC, PLR and NLR were not found to be very significantly associated with mortality.

**Table 7:** Model 1.

| Class | Precision | Recall | f1-score | support |
| --- | --- | --- | --- | --- |
| 1 (Survived) | 0.93 | 0.89 | 0.91 | 152 |
| 2 (Did not survive) | 0.71 | 0.78 | 0.75 | 51 |
| Accuracy |  |  | 0.87 | 203 |

**Table 8:** Multivariate logistic regression analysis model.

|  |  |  |  |
| --- | --- | --- | --- |
| Dep. Variable: | DF.iloc[:, 4] | No. Observations: | 203 |
| Model: | Logit | Df Residuals: | 198 |
| Method: | MLE | Df Model: | 4 |
| Date: | Thu, 15 Apr 2021 | Pseudo R-squ.: | 0.5400 |
| Time: | 12:26:58 | Log-Likelihood: | -52.641 |
| converged: | True | LL-Null: | -114.43 |
| Covariance Type: | nonrobust | LLR p-value: | 9.210e-26 |

|  | coef | std err | z | P>\|z\| | [0.025 | 0.975] |
| --- | --- | --- | --- | --- | --- | --- |
| Intercept | 8.2843 | 1.445 | 5.732 | 0.000 | 5.451 | 11.117 |
| DF.iloc[:, 0] | -0.0002 | 5.39e-05 | -3.747 | 0.000 | -0.000 | -9.64e-05 |
| DF.iloc[:, 1] | 0.0004 | 0.000 | 1.580 | 0.114 | -0.000 | 0.001 |
| DF.iloc[:, 2] | 0.0013 | 0.002 | 0.595 | 0.552 | -0.003 | 0.006 |
| DF.iloc[:, 3] | -2.7171 | 0.464 | -5.852 | 0.000 | -3.627 | -1.807 |

**Figure 1:**
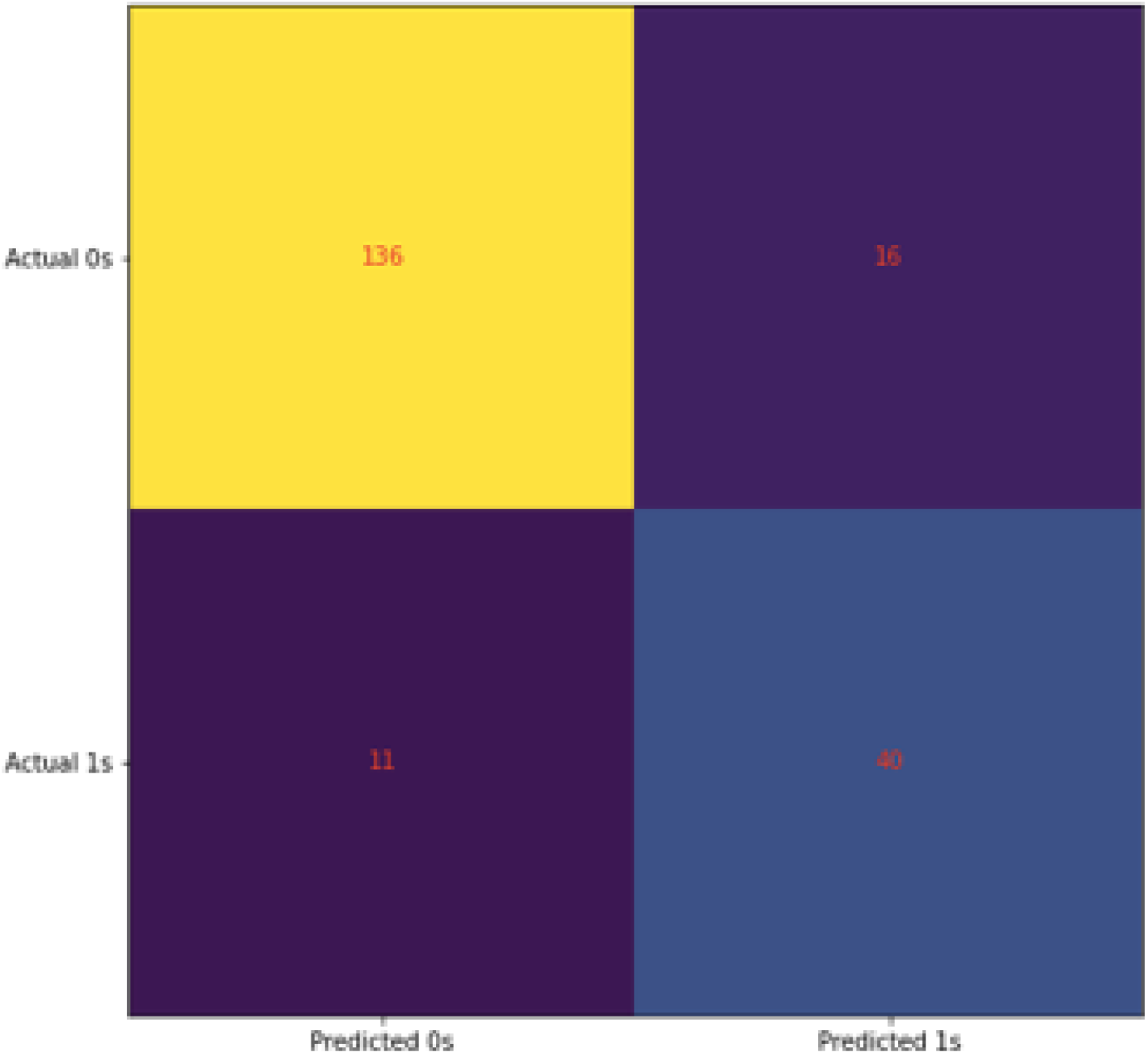
Confusion matrix for model 1

Certain variables used in this model showed high degree of correlation and therefore redundancy. A second model was built using multivariate logistic regression in Python 3.7. The variables ANC, ALC, PLR and severity were found to be independent of one another. NLR was redundant because both ALC and ANC were used. TLC was redundant because if 0.97 correlation coefficient with ANC. The data was first scaled and then a model was built. The default cut off of 0.5 was retained for both models.

**Table 9:** Model 2

| Logit Regression Results |  |  |  |  |  |  |
| --- | --- | --- | --- | --- | --- | --- |
| Dep. Variable: | Survival | No. Observations: | 203 |  |  |  |
| Model: | Logit | Df Residuals: | 199 |  |  |  |
| Method: | MLE | Df Model: | 3 |  |  |  |
| Date: | Thu, 15 Apr 2021 | Pseudo R-squ.: | 0.1676 |  |  |  |
| Time: | 13:10:19 | Log-Likelihood: | -95.249 |  |  |  |
| converged: | True | LL-Null: | -114.43 |  |  |  |
| Covariance Type: | nonrobust | LLR p-value: | 2.374e-08 |  |  |  |
|  | coef | std err | z | P>\|z\| | [0.025 | 0.975] |
| x1 | -1.0480 | 0.288 | -3.639 | 0.000 | -1.612 | -0.484 |
| x2 | 0.2219 | 0.256 | 0.867 | 0.386 | -0.280 | 0.723 |
| x3 | 0.1740 | 0.238 | 0.731 | 0.465 | -0.292 | 0.640 |
| x4 | -1.1087 | 0.207 | -5.363 | 0.000 | -1.514 | -0.704 |

**Figure 2:**
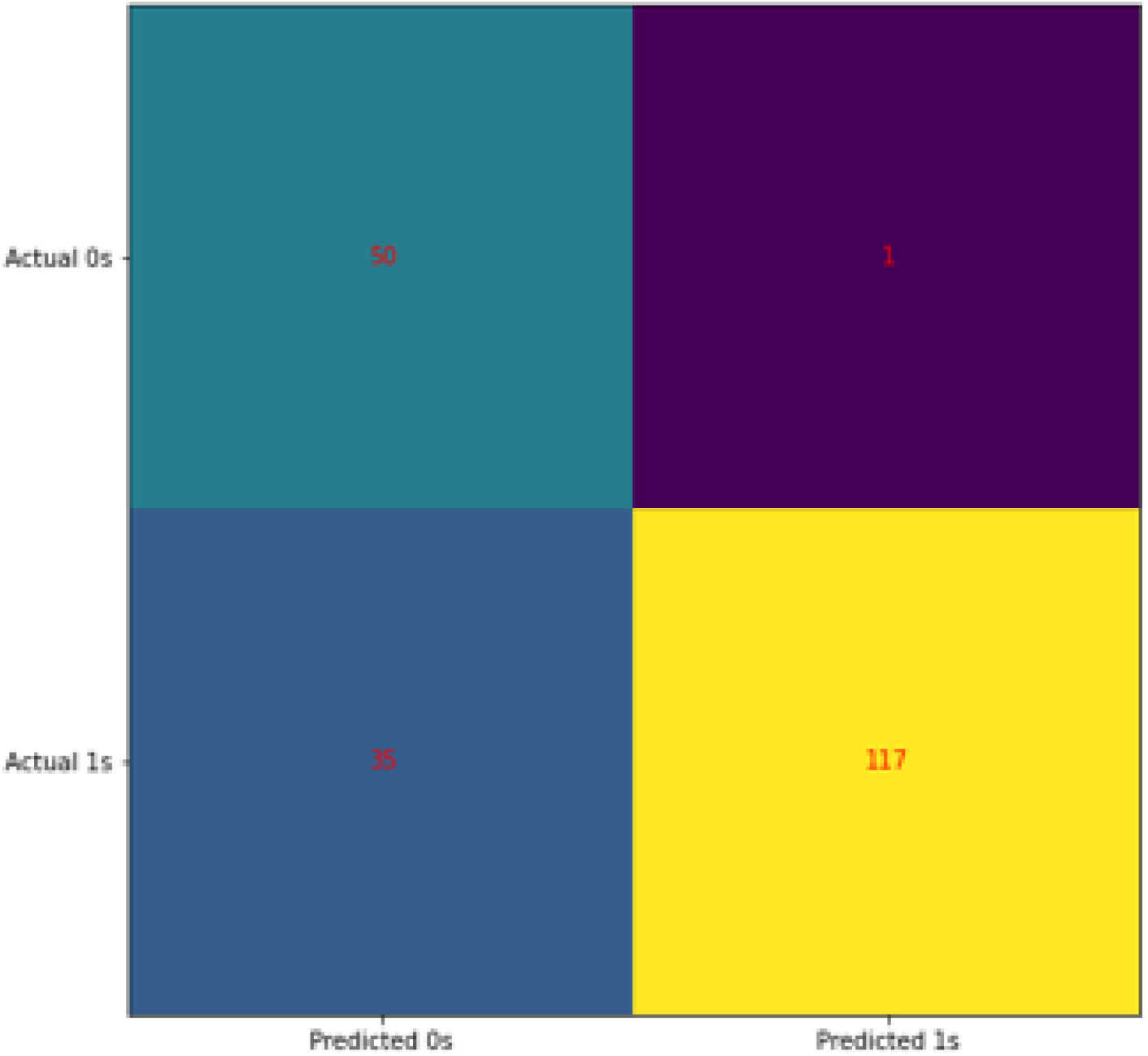
Confusion matrix for model 2

Again the association of ANC and severity with survival was significant with p values approaching zero, whereas ALC and PLR were not significantly associated. Although the overall accuracy of the second model was lower with an accuracy score of 82%, it had a very high sensitivity for predicting mortality (98%). This demonstrated that a combination of severity and ANC at presentation (start of pneumonia) have a high sensitivity for predicting adverse outcomes in COVID. Given these findings a third model was built using only clinical severity (determined by SpO2) and ANC as the predictor variables. This model performed just as well as the second one with regards to mortality prediction.

**Table 10:**
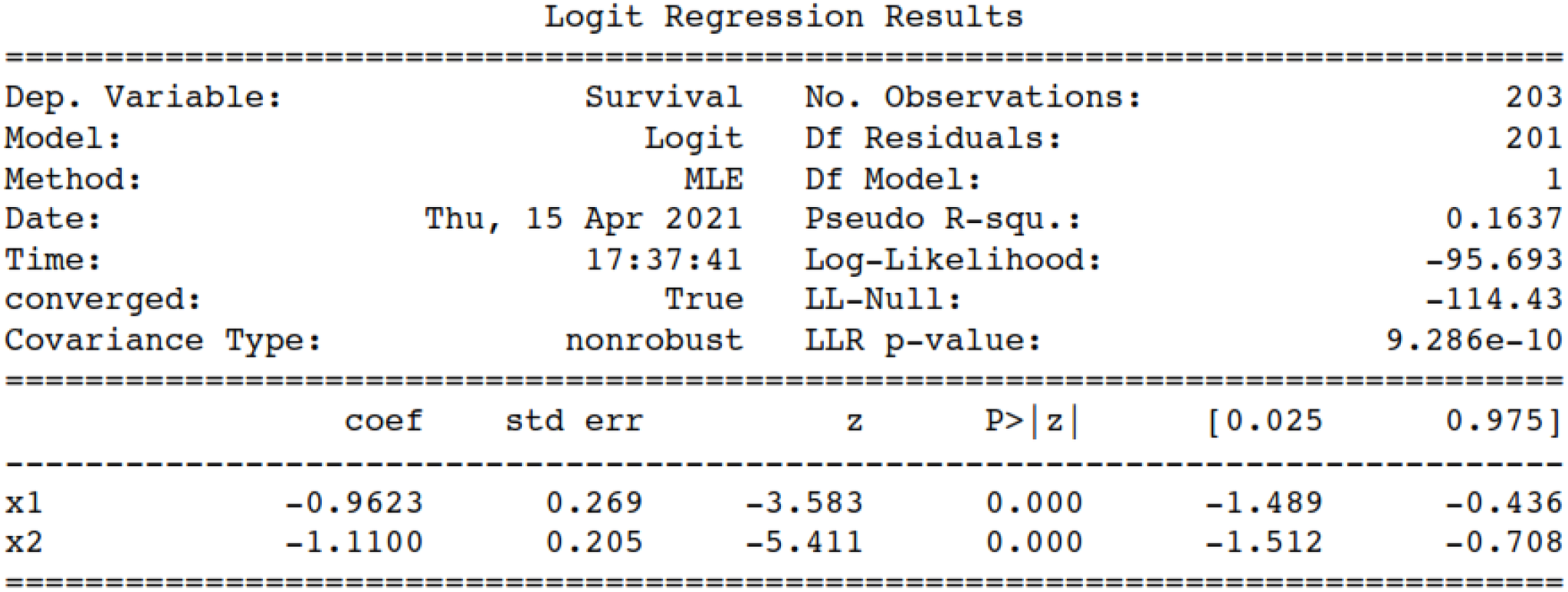
Model 3

**Figure 3:**
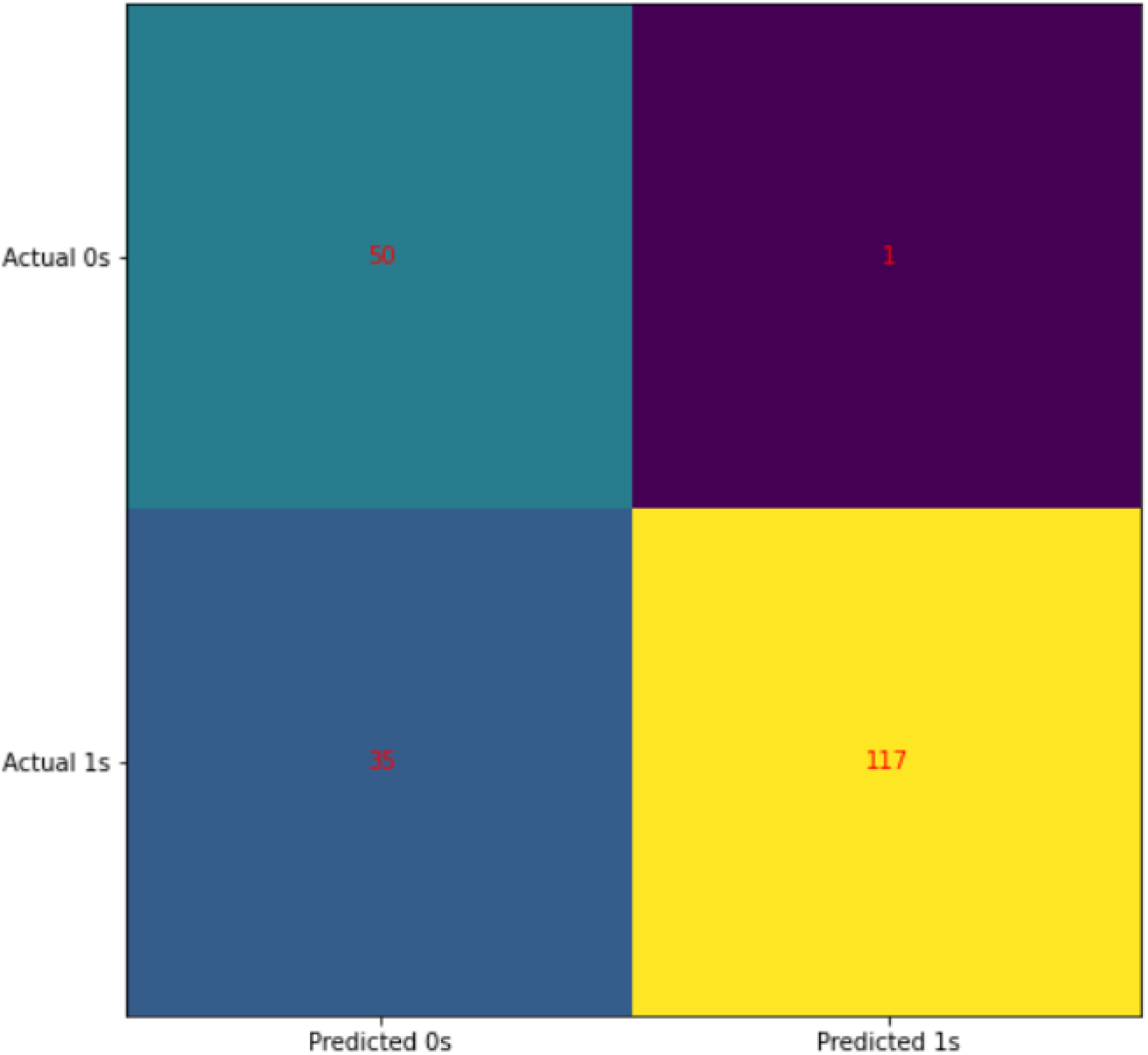
Confusion matrix for model 3

Since ANC and TLC correlated highly with one another, we tested the model efficacy with TLC used in lieu of ANC and found the result to be the same. On this basis we also recommend the use of TLC in place of ANC in situations where an accurate differential count may not be available. The model, developed in Python 3.7, is available for online use at the IP address https://mybinder.org/v2/gh/jungleeToofan/covidModel.git/HEAD?urlpath=%2Fvoila%2Frender%2FCovidModel.ipynb. We have named this model the 3C-M score (Clinical severity & blood Counts in Covid-19 pneumonia for Mortality prediction). It is a binary classification tool, with values greater than 0.5 signifying increased risk of mortality. The higher the score, the more accurate the classifiers performance in predicting mortality.

Summary of key findings in our study:

1. Absolute neutrophil count is an independent, early, strong predictor of both disease severity and survival. During early disease course it correlates better than lymphocyte count and NLR with severity and survival.
2. The NLR means even on Day 1 were significantly higher in those who did not survive. The rise in NLR at the onset of pneumonia was primarily driven by a higher neutrophil count rather than a low lymphocyte count.
3. Early lymphocyte count correlates poorly with survival and severity.
4. It is possible to do an early triage on the basis of simply and widely available clinical and laboratory information. This might be of particular benefit in aiding decision making in a primary healthcare setting regarding rapid escalation of care in those who need it. The 3C-M model was developed for use in such resource limited settings for timely estimation of mortality risk.

## Discussion

In our study we found a strong association of neutrophilic leukocytosis with severity as well as survival in COVID-19 pneumonia. The mean neutrophil count in those who survived was 6253.361 and those who didn’t were 13665.92. The difference was highly significant with a p value of 0.001. The table below includes a list of studies which have also found ANC to be significantly associated with survival and severity. Another recent study by Wei et al evaluated neutrophil count as a predictor variable(23). They studied the correlation of baseline neutrophil count (bNC) and neutrophil change rate (NCR) with death. They concluded that bNC had a U-shaped association with mortality; lower counts were associated with improved survival and higher counts were associated with mortality. The NRC could not help in risk stratification of patients.

Another recently published study found that total white cell count was significantly associated with severity, but lymphocyte count was not(24). This demonstrates that a new understanding of COVID-19 pneumonia pathogenesis is emerging; one which places neutrophils front and centre as the major effector cells in the disease process. It also shows that neutrophil count can be useful in developing predictive models of COVID-19 especially in a resource limited setting.

It is important to note that there are other, non-COVID related factors which can influence the neutrophil count and thereby reduce their relevance as a prognostic marker. Presence of a secondary bacterial infection and the initiation of steroids are two such examples.

Whether this parameter would retain value in these situations is difficult to say. At present is prudent to stress the importance of using neutrophil count obtained at the onset of respiratory complications for assessment of severity and triage.

| S. No. | Study | Area | Size | TLC Survival (p) | Severity (p) | ANC Survival (p) | Severity (p) |
| --- | --- | --- | --- | --- | --- | --- | --- |
| 1 | Huang et al(25) | China | 41 | NR | 0.011 | NR | 0.00069 |
| 2 | Liu et al(26) | China | 12 | NR | NR | NR | 0.018 |
| 3 | Qian et al(27) | China | 91 | NR | 0.01 | NR | 0.00041 |
| 4 | Wang et al(4) | China | 138 | NR | 0.003 | NR | <0.001 |
| 5 | Wang et al(28) | China | 69 | NR | 0.006 | NR | <0.001 |
| 6 | Wu et al(29) | China | 197 | NR | <0.001 | 0.03 | <0.001 |
| 8 | Xiong et al(30) | China | 116 | NR | <0.001 | NR | <0.001 |
| 9 | Liu et al(26) | China | 155 | NR | NR | NR | 0.017 |
| 10 | Yang et al(31) | China | 61 | NR | 0.215 | NR | 0.025 |
| 11 | Li et al(32) | China | 93 | NR | 0.006 | NR | <0.001 |
| 13 | Shi et al(24) | China | 315 | <0.0001 | NR | <0.0001 | NR |
| 14 | Guan et al(6) | China | 1099 | NR | <0.0001 | NR | 0.002 |
| 15 | Our study | India | 203 | 0.001 | 0.001 | 0.001 | 0.001 |

The earlier studies reported that leukopenia and lymphopaenia were the predominant hematological findings associated with COVID-19 pneumonia. The table below lists the studies which reported information regarding lymphopaenia in COVID-19. The results were not consistent with the incidence of lymphopaenia varying from 22% to 83% in various studies. Some of these studies also compared the lymphocyte count of survivors with non-survivors. The largest by Guan et al, with a sample size of 1099, found the incidence of lymphopaenia to be 92.6% in those with severe disease compared to 82.5% in those with non severe disease(6). Studies by Yang and Zhou concluded that lymphocyte count on admission were not significantly different between survivors and non-survivors(31), (15). Studies by Fan and Tan showed that lymphocyte count as a variable for predicting severity is significant only if serial values are obtained and there is a persistent fall over the course of illness(33), (16). Wu et al concluded that lower lymphocyte count was associated with development of ARDS but not with survival in those who had developed ARDS(5). In our study we found that the mean lymphocyte count on day of admission has significance in predicting severity and survival only on univariate analysis. On multivariate analysis this significance is lost. All these highlight that lymphocyte count is not a robust variable for purpose of initial triage.

| S. No. | Study | Area | Size | Low TLC (%) | Low ALC (%) | Increased NLR >3.3 (%) |
| --- | --- | --- | --- | --- | --- | --- |
| 1 | Chen et al(3) | China | 29 | 21 | 69 | NR |
| 2 | Fan et al(33) | Singapore | 65 | 29.2 | 36.9 | NR |
| 3 | Guan et al(6) | China | 1099 | 33.7 | 83.2 | NR |
| 4 | Huang et al(25) | China | 41 | 25 | 63 | NR |
| 5 | Liu et al(26) | China | 30 | 26.6 | 40 | NR |
| 6 | Qian et al(27) | China | 91 | 15.3 | 30.7 | NR |
| 7 | Wang et al(4) | China | 69 | 54 | 42 | NR |
| 8 | Zhang et al(34) | China | 140 | 19.6 | 75.4 | NR |
| 9 | Zhou et al(15) | China | 140 | 17 | 40 | NR |
| 10 | Arentz et al(13) | USA | 21 | NR | 67 | NR |
| 11 | Bhatraju et al(35) | USA | 24 | 4 | 75 | NR |
| 12 | Chen et al(3) | China | 99 | 9 | 35 | NR |
| 13 | Wang et al(4) | China | 60 | 32 | 72 | NR |
| 14 | Shi et al(36) | China | 635 | 12 | 30 | NR |
| 15 | Chen et al(37) | China | 291 | 36.1 | 22.7 | 26.8 |
| 16 | Our study | India | 203 | 5.41 | 26.2 | 65.02 |

Another parameter, the neutrophil to lymphocyte ratio (NLR), also has positive correlation with mortality. The mean NLR was significantly different in those who survived (5.3) versus those who did not survive (12.5) with a p value approaching 0.001. With regard to severity the difference in the mean NLR was only significant in those with clinically mild (4.8) and severe (10.0) pneumonia; it could not differentiate between the mild-moderate and moderate-severe categories with confidence. However, a multivariate analysis showed us that the higher NLR at baseline was driven by a higher neutrophil count confounding the relevance of NLR as an independent predictor. A comparison of absolute neutrophil count and NLR showed that neutrophil count performed better as predictor variable. This brought to the fore the issue of redundancy among variables and the need to ensure that one is vigilant about keeping it to a minimum. So far an entire basket of parameters has been placed at the physician’s disposal to pick and choose from. This can and does lead to an unfortunate loss of clarity.

Our research raises an important question - why does early neutrophilic leukocytosis correlate with severity and survival? There has been a recent surge of interest in evaluating the role of neutrophil driven inflammation as an important contributory factor for the cytokine storm observed in COVID-19 as well as in other viral infections(38). A recent gene network analysis found that infected cells in the lung express neutrophil attracting chemokine and BAL fluid analysis also showed an up regulation of neutrophil genes and chemokine(39), (40), (41), (42). Neutrophil extracellular trap (NET) generation has been the focus of intense study in the past decade or so(43), (44). Several studies have implicated SARS CoV 2 driven NETs as an important mediator of inflammation in COVID-19, with signatures of neutrophil activation consistently associated with inferior outcomes. A recent study specifically implicates NETs in driving immune thrombosis in COVID-19. A comparison between platelet activation and neutrophil activation in those who develop over thromboembolism in COVID-19 showed that neutrophil activation was more significantly associated with the same(45). Another studied identified D-Dimer levels and neutrophil count as biomarkers which can be potential predictive markers for pulmonary embolism(46). Research is also indicative of the role of persistent innate immune activation in hyper-inflammatory states(47), (48), (49), (50). The exact immune-cytokine subset which proceeds towards the development of a lung inflammation syndrome has not yet been delineated. Indeed there may not even be any ‘one’ immune signature of this state. This is an area which requires extensive research but one can cautiously state that neutrophils might hold one of the keys to unlocking this mystery.

The second question we attempted to tackle was building a simple model for the purpose of triage in a resource limited setting based on clinical and basic laboratory information that is the CBC. Two multivariate logistic regression models were built using all the data available to us. While one had a good overall precision of 87%, the accuracy for identifying survivors was 93% but for non survivors it was only 71%. A second model was built after excluding redundant variables; ANC, ALC, PLR and clinical severity were included in the second one. This model gave nearly equal weightage to ANC and clinical severity. Though the overall accuracy was lower at 82%, it showed remarkably greater sensitivity for mortality prediction accurately predicting 50 of 51 deaths. The model was made available for online use as it requires a minimal of information which can be quickly and easily available even in primary health care setting. This model has not yet been validated externally; it is currently undergoing a second, internal, retrospective validation at our centre.

It is pertinent to mention that many other models have also been built which attempt to predict mortality in COVID-19. The simple reason for adding yet another one to the plethora already available is the ease of use and the minimal information required. However potential drawbacks remain, a major one being the time of applicability. Our model was used on patients who presented to the hospital with onset of pneumonia symptoms, however it cannot be assumed that all patients will present at a similar time in their disease course.

A recent study compared the performance of eleven prediction models with MICU admission and mortality as independent and composite end points(51). Of these, the RISE UP and 4C models performed decently with an AUC of 0.83 and 0.84 for predicting 30 day mortality respectively(52), (53). A recent critical analysis has cautiously endorsed the 4C score for guidance of decision making(53). They have also, simultaneously, highlighted the potential limitations of this score like the ‘treatment paradox’. As the disease continues to evolve and our understanding of it continues to change, the impact assessment of these models will require rigorous and continued assessment in real time in order to retain validity. Keeping these relevant critiques in mind we decided to wait to prospectively evaluate our model before recommending it for general use.

## Conclusion

On the basis of our study and the review of literature we would like to stress the significance of an early pronounced neutrophilic response as a sensitive predictor of disease severity and outcome. This knowledge can be particularly useful in a resource limited setting. A combination of clinical severity and absolute neutrophil count has allowed us to build a model, which can help the treating doctor to sensitively pick up those at risk of poor outcome at presentation; and this can potentially drive early intervention. The pathological role of high neutrophil count in driving lung inflammation in COVID-19 needs to be studied in further detail.

## Data Availability

Data is available for perusal from the department of Medicine, Lady Hardinge Medical college, Delhi, India in physical format since the college does not yet have an electronic records department.

## Notes

### Competing Interest Statement

The authors have declared no competing interest.

### Funding Statement

No external funding recieved for this project

### Author Declarations

Ethics Committee- L.H.M.C, Delhi, India

## References

1. Zhu N, Zhang D, Wang W, Li X, Yang B, Song J, et al. A Novel Coronavirus from Patients with Pneumonia in China, 2019. N Engl J Med. 2020 Feb 20;382(8):727–33.

2. Ksiazek TG, Erdman D, Goldsmith CS, Zaki SR, Peret T, Emery S, et al. A novel coronavirus associated with severe acute respiratory syndrome. N Engl J Med. 2003 May 15;348(20):1953–66.

3. Chen N, Zhou M, Dong X, Qu J, Gong F, Han Y, et al. Epidemiological and clinical characteristics of 99 cases of 2019 novel coronavirus pneumonia in Wuhan, China: a descriptive study. Lancet Lond Engl. 2020 Feb 15;395(10223):507–13.

4. Wang D, Hu B, Hu C, Zhu F, Liu X, Zhang J, et al. Clinical Characteristics of 138 Hospitalized Patients With 2019 Novel Coronavirus-Infected Pneumonia in Wuhan, China. JAMA. 2020 Mar 17;323(11):1061–9.

5. Wu Z, McGoogan JM. Characteristics of and Important Lessons From the Coronavirus Disease 2019 (COVID-19) Outbreak in China: Summary of a Report of 72 314 Cases From the Chinese Center for Disease Control and Prevention. JAMA. 2020 Apr 7;323(13):1239–42.

6. Guan W-J, Ni Z-Y, Hu Y, Liang W-H, Ou C-Q, He J-X, et al. Clinical Characteristics of Coronavirus Disease 2019 in China. N Engl J Med. 2020 Apr 30;382(18):1708–20.

7. Mehta P, McAuley DF, Brown M, Sanchez E, Tattersall RS, Manson JJ, et al. COVID-19: consider cytokine storm syndromes and immunosuppression. Lancet Lond Engl. 2020 Mar 28;395(10229):1033–4.

8. Yuki K, Fujiogi M, Koutsogiannaki S. COVID-19 pathophysiology: A review. Clin Immunol Orlando Fla. 2020 Jun;215:108427.

9. Tang Y, Liu J, Zhang D, Xu Z, Ji J, Wen C. Cytokine Storm in COVID-19: The Current Evidence and Treatment Strategies. Front Immunol. 2020;11:1708.

10. Qin C, Zhou L, Hu Z, Zhang S, Yang S, Tao Y, et al. Dysregulation of Immune Response in Patients With Coronavirus 2019 (COVID-19) in Wuhan, China. Clin Infect Dis Off Publ Infect Dis Soc Am. 2020 Jul 28;71(15):762–8.

11. Coomes EA, Haghbayan H. Interleukin-6 in Covid-19: A systematic review and meta-analysis. Rev Med Virol. 2020 Nov;30(6):1–9.

12. Magro G. SARS-CoV-2 and COVID-19: Is interleukin-6 (IL-6) the “culprit lesion” of ARDS onset? What is there besides Tocilizumab? SGP130Fc. Cytokine X. 2020 Jun;2(2):100029.

13. Arentz M, Yim E, Klaff L, Lokhandwala S, Riedo FX, Chong M, et al. Characteristics and Outcomes of 21 Critically Ill Patients With COVID-19 in Washington State. JAMA. 2020 Apr 28;323(16):1612–4.

14. Deng Y, Liu W, Liu K, Fang Y-Y, Shang J, Zhou L, et al. Clinical characteristics of fatal and recovered cases of coronavirus disease 2019 in Wuhan, China: a retrospective study. Chin Med J (Engl). 2020 Jun 5;133(11):1261–7.

15. Zhou F, Yu T, Du R, Fan G, Liu Y, Liu Z, et al. Clinical course and risk factors for mortality of adult inpatients with COVID-19 in Wuhan, China: a retrospective cohort study. Lancet Lond Engl. 2020 Mar 28;395(10229):1054–62.

16. Tan L, Wang Q, Zhang D, Ding J, Huang Q, Tang Y-Q, et al. Lymphopenia predicts disease severity of COVID-19: a descriptive and predictive study. Signal Transduct Target Ther. 2020 Mar 27;5(1):33.

17. Lippi G, Favaloro EJ. D-dimer is Associated with Severity of Coronavirus Disease 2019: A Pooled Analysis. Thromb Haemost. 2020 May;120(5):876–8.

18. Lillicrap D. Disseminated intravascular coagulation in patients with 2019-nCoV pneumonia. J Thromb Haemost JTH. 2020 Apr;18(4):786–7.

19. Lippi G, Plebani M. Procalcitonin in patients with severe coronavirus disease 2019 (COVID-19): A meta-analysis. Clin Chim Acta Int J Clin Chem. 2020 Jun;505:190–1.

20. Han H, Yang L, Liu R, Liu F, Wu K-L, Li J, et al. Prominent changes in blood coagulation of patients with SARS-CoV-2 infection. Clin Chem Lab Med. 2020 Jun 25;58(7):1116–20.

21. Cheng L, Li H, Li L, Liu C, Yan S, Chen H, et al. Ferritin in the coronavirus disease 2019 (COVID-19): A systematic review and meta-analysis. J Clin Lab Anal. 2020 Oct;34(10):e23618.

22. Carubbi F, Salvati L, Alunno A, Maggi F, Borghi E, Mariani R, et al. Ferritin is associated with the severity of lung involvement but not with worse prognosis in patients with COVID-19: data from two Italian COVID-19 units. Sci Rep. 2021 Mar 1;11(1):4863.

23. Fu W, Chen C, Chen X-L, Wang K, Zuo P, Liu Y, et al. A U-shaped association between baseline neutrophil count and COVID-19-related mortality: A retrospective cohort study. J Med Virol. 2021 Jul;93(7):4265–72.

24. Shi L, Wang Y, Liang X, Xiao W, Duan G, Yang H, et al. Is neutrophilia associated with mortality in COVID-19 patients? A meta-analysis and meta-regression. Int J Lab Hematol. 2020 Dec;42(6):e244–7.

25. Huang C, Wang Y, Li X, Ren L, Zhao J, Hu Y, et al. Clinical features of patients infected with 2019 novel coronavirus in Wuhan, China. Lancet Lond Engl. 2020 Feb 15;395(10223):497–506.

26. Liu K, Chen Y, Lin R, Han K. Clinical features of COVID-19 in elderly patients: A comparison with young and middle-aged patients. J Infect. 2020 Jun;80(6):e14–8.

27. Qian G-Q, Yang N-B, Ding F, Ma AHY, Wang Z-Y, Shen Y-F, et al. Epidemiologic and clinical characteristics of 91 hospitalized patients with COVID-19 in Zhejiang, China: a retrospective, multi-centre case series. QJM Mon J Assoc Physicians. 2020 Jul 1;113(7):474–81.

28. Wang Z, Yang B, Li Q, Wen L, Zhang R. Clinical Features of 69 Cases With Coronavirus Disease 2019 in Wuhan, China. Clin Infect Dis Off Publ Infect Dis Soc Am. 2020 Jul 28;71(15):769–77.

29. Wu C, Chen X, Cai Y, Xia J, Zhou X, Xu S, et al. Risk Factors Associated With Acute Respiratory Distress Syndrome and Death in Patients With Coronavirus Disease 2019 Pneumonia in Wuhan, China. JAMA Intern Med. 2020 Jul 1;180(7):934–43.

30. Xiong S, Liu L, Lin F, Shi J, Han L, Liu H, et al. Clinical characteristics of 116 hospitalized patients with COVID-19 in Wuhan, China: a single-centered, retrospective, observational study. BMC Infect Dis. 2020 Oct 22;20(1):787.

31. Yang X, Yu Y, Xu J, Shu H, Xia J, Liu H, et al. Clinical course and outcomes of critically ill patients with SARS-CoV-2 pneumonia in Wuhan, China: a single-centered, retrospective, observational study. Lancet Respir Med. 2020 May;8(5):475–81.

32. Li L, Yang L, Gui S, Pan F, Ye T, Liang B, et al. Association of clinical and radiographic findings with the outcomes of 93 patients with COVID-19 in Wuhan, China. Theranostics. 2020;10(14):6113–21.

33. Fan BE, Chong VCL, Chan SSW, Lim GH, Lim KGE, Tan GB, et al. Hematologic parameters in patients with COVID-19 infection. Am J Hematol. 2020 Jun;95(6):E131–4.

34. Zhang J-J, Dong X, Cao Y-Y, Yuan Y-D, Yang Y-B, Yan Y-Q, et al. Clinical characteristics of 140 patients infected with SARS-CoV-2 in Wuhan, China. Allergy. 2020 Jul;75(7):1730–41.

35. Bhatraju PK, Ghassemieh BJ, Nichols M, Kim R, Jerome KR, Nalla AK, et al. Covid-19 in Critically Ill Patients in the Seattle Region - Case Series. N Engl J Med. 2020 May 21;382(21):2012–22.

36. Shi Q, Zhang X, Jiang F, Zhang X, Hu N, Bimu C, et al. Clinical Characteristics and Risk Factors for Mortality of COVID-19 Patients With Diabetes in Wuhan, China: A Two-Center, Retrospective Study. Diabetes Care. 2020 Jul;43(7):1382–91.

37. Chen X, Zheng F, Qing Y, Ding S, Yang D, Lei C, et al. Epidemiological and clinical features of 291 cases with coronavirus disease 2019 in areas adjacent to Hubei, China: a double-center observational study. medRxiv. 2020 Jan 1;2020.03.03.20030353.

38. Wang J, Jiang M, Chen X, Montaner LJ. Cytokine storm and leukocyte changes in mild versus severe SARS-CoV-2 infection: Review of 3939 COVID-19 patients in China and emerging pathogenesis and therapy concepts. J Leukoc Biol. 2020 Jul;108(1):17–41.

39. Xiong Y, Liu Y, Cao L, Wang D, Guo M, Jiang A, et al. Transcriptomic characteristics of bronchoalveolar lavage fluid and peripheral blood mononuclear cells in COVID-19 patients. Emerg Microbes Infect. 2020 Dec;9(1):761–70.

40. Didangelos A. COVID-19 Hyperinflammation: What about Neutrophils? mSphere. 2020 Jun 24;5(3):e00367–20.

41. Zhou Z, Ren L, Zhang L, Zhong J, Xiao Y, Jia Z, et al. Heightened Innate Immune Responses in the Respiratory Tract of COVID-19 Patients. Cell Host Microbe. 2020 Jun 10;27(6):883-890.e2.

42. Ronit A, Berg RMG, Bay JT, Haugaard AK, Ahlström MG, Burgdorf KS, et al. Compartmental immunophenotyping in COVID-19 ARDS: A case series. J Allergy Clin Immunol. 2021 Jan;147(1):81–91.

43. Wang J, Li Q, Yin Y, Zhang Y, Cao Y, Lin X, et al. Excessive Neutrophils and Neutrophil Extracellular Traps in COVID-19. Front Immunol. 2020;11:2063.

44. Mozzini C, Girelli D. The role of Neutrophil Extracellular Traps in Covid-19: Only an hypothesis or a potential new field of research? Thromb Res. 2020 Jul;191:26–7.

45. Zuo Y, Zuo M, Yalavarthi S, Gockman K, Madison JA, Shi H, et al. Neutrophil extracellular traps and thrombosis in COVID-19. J Thromb Thrombolysis. 2021 Feb;51(2):446–53.

46. Thoreau B, Galland J, Delrue M, Neuwirth M, Stepanian A, Chauvin A, et al. D-Dimer Level and Neutrophils Count as Predictive and Prognostic Factors of Pulmonary Embolism in Severe Non-ICU COVID-19 Patients. Viruses. 2021 Apr 26;13(5):758.

47. Li G, Fan Y, Lai Y, Han T, Li Z, Zhou P, et al. Coronavirus infections and immune responses. J Med Virol. 2020 Apr;92(4):424–32.

48. Blanco-Melo D, Nilsson-Payant BE, Liu W-C, Uhl S, Hoagland D, Møller R, et al. Imbalanced Host Response to SARS-CoV-2 Drives Development of COVID-19. Cell. 2020 May 28;181(5):1036-1045.e9.

49. Rodrigues TS, de Sá KSG, Ishimoto AY, Becerra A, Oliveira S, Almeida L, et al. Inflammasomes are activated in response to SARS-CoV-2 infection and are associated with COVID-19 severity in patients. J Exp Med. 2021 Mar 1;218(3):e20201707.

50. Freeman TL, Swartz TH. Targeting the NLRP3 Inflammasome in Severe COVID-19. Front Immunol. 2020;11:1518.

51. van Dam PMEL, Zelis N, van Kuijk SMJ, Linkens AEMJH, Brüggemann Rag, Spaetgens B, et al. Performance of prediction models for short-term outcome in COVID-19 patients in the emergency department: a retrospective study. Ann Med. 2021 Dec;53(1):402–9.

52. Knight SR, Ho A, Pius R, Buchan I, Carson G, Drake TM, et al. Risk stratification of patients admitted to hospital with covid-19 using the ISARIC WHO Clinical Characterisation Protocol: development and validation of the 4C Mortality Score. BMJ. 2020 Sep 9;370:m3339.

53. Wellbelove Z, Walsh C, Perinpanathan T, Lillie P, Barlow G. Comparing the 4C mortality score for COVID-19 to established scores (CURB65, CRB65, qSOFA, NEWS) for respiratory infection patients. J Infect. 2021 Mar;82(3):414–51.

